# Comparison of carbon dioxide control during pressure controlled versus pressure regulated volume controlled ventilation in children (CoCO2): protocol for a pilot digital randomized controlled trial

**DOI:** 10.1101/2024.03.29.24305023

**Authors:** Rebeca Mozun, Daphné Chopard, Florian Zapf, Philipp Baumann, Barbara Brotschi, Anika Adam, Vera Jaeggi, Beat Bangerter, Kristen Gibbons, Juerg Burren, Luregn J Schlapbach

## Abstract

**Introduction:** Digital trials are a promising strategy to increase the evidence base for common interventions and may convey considerable efficiency benefits in trial conduct. Although paediatric intensive care units (PICUs) are rich in routine electronic data, highly pragmatic digital trials in this field remain scarce. There are unmet evidence needs for optimal mechanical ventilation modes in paediatric intensive care. We aim to test the feasibility of a digital PICU trial comparing two modes of invasive mechanical ventilation using carbon dioxide (CO_2_) control as the outcome measure.

**Methods and analysis:** Single-centre, open-labelled, randomized controlled pilot trial with two parallel treatment arms comparing pressure control (PC) vs pressure-regulated volume control (PRVC). Patients are eligible if aged <18 years, weighing >2 kg, have an arterial line, and require >60 minutes of mechanical ventilation during PICU hospitalization at the University Children’s Hospital Zurich. Exclusion criteria include cardiac shunt lesions, pulmonary hypertension under treatment, and intracranial hypertension. CO_2_ is measured using three methods: end-tidal (continuous), transcutaneous (continuous), and blood gas analyses (intermittent). Baseline, intervention, and outcome data are collected electronically from the patients’ routine electronic health records. The primary feasibility outcome is adherence to the assigned ventilation mode, while the primary physiological outcome is the proportion of time spent within the target range of CO_2_ (end-tidal, normocarbia defined as CO_2_ ≥ 4.5, ≤ 6 kPa). Both outcomes are captured digitally every minute from randomization until censoring (48 hours after randomization, extubation, discharge, or death, whichever comes first). Analysis will occur on an intention-to-treat basis. We aim to enrol 60 patients in total. Recruitment started in January 2024 and is planned to continue for 6 months.

**Ethics and dissemination:** This study received ethical approval (BASEC 2022-00829). Study results will be disseminated through publication in a peer-reviewed journal and other media like podcasts.

**Trial registration number:** NCT058431

**ARTICLE SUMMARY:**

- This study compares two commonly used modes of invasive mechanical ventilation in a randomized design. The trial will provide feasibility data to inform the conduct of digital trials by using electronic patient data directly extracted from the source systems, minimizing manual data collection and associated bias and thereby increasing local readiness for more efficient clinical trial conduct.
- Outcomes of this pilot trial relate to feasibility and physiological measures; future larger trials should also explore patient-centred outcomes.
- Blinding is not possible due to the nature of the intervention.
- Technical issues that may affect the availability or accuracy of data may arise and will be documented.
- Some aspects of digital trials, such as electronic informed consent, are not implemented in this trial.

## INTRODUCTION

Trials using digital recordings of routine clinical data have great potential to decrease the resources required to conduct clinical trials, which remain one of the major obstacles to faster evidence generation in clinical care. The COVID pandemic has demonstrated the need for rapid implementation of clinical trials in intensive care, particularly including platform or adaptive designs^1,2^. Advantages of digital clinical trials include facilitation of recruitment, optimization of prospective data collection, workflows, and monitoring, as well as innovative methods to automate processes, make predictions, or assist in phenotyping and result interpretation^3^. Intensive care units (ICUs) represent an attractive setting for digital trials given the rich data environment with high temporal resolution^4^.

Approximately one in three children receives invasive mechanical ventilation when admitted to PICU^5^. Invasive mechanical ventilation can save lives while bearing risks of ventilator-induced lung injury (VILI)^6^. Other negative aspects directly related to invasive mechanical ventilation include ventilator-associated events such as ventilator-associated pneumonia, and pneumothorax, side effects of sedation such as neurotoxicity, withdrawal, delirium, and family burden due to disruptive interactions. Mechanical ventilation’s main goal is to provide appropriate gas exchange. Suboptimal mechanical ventilation may lead to fluctuations in carbon dioxide (CO_2_) with associated changes in cerebral and pulmonary blood flow^7^. These changes can aggravate cerebral ischemia or swelling and pulmonary arterial hypertension. Prolonged intubation and delayed weaning increase the risk of chronic lung disease, prolonged PICU stay, critical illness neuromyopathy, and worse patient-centred outcomes^8^.

It is not known which ventilation mode is most effective with the least harm in children, despite recent technological advances in paediatric mechanical ventilation and the availability of digital ventilation monitoring data^9^. Classical ventilation modes include conventional pressure-controlled (PC) and volume-controlled (VC) modes. Modern ventilators offer algorithm-based adaptive ventilation modes to maximize lung protection^10^. Pressure-regulated volume control (PRVC) is one such adaptive mode that aims to achieve optimal oxygenation and lung recruitment at lower mean airway pressures while maintaining a defined tidal volume (TV)^7^. These adaptive modes rapidly adjust to changes in lung compliance, thereby reducing lung injury, providing a stable minute volume, and stabilizing gas exchange. In a Cochrane review, neonates (mostly preterm or low birth weight) ventilated with PRVC modes had lower rates of death or bronchopulmonary dysplasia, pneumothorax, hypocarbia, severe cranial ultrasound pathologies, and duration of ventilation compared with infants ventilated with PC modes^11^. However, there is insufficient evidence to determine the best ventilation mode in critically ill children, and clinicians’ choice is often based on personal or institutional preference^9,12^.

### Aims and hypothesis

The aims of this study are to test the feasibility of a digital trial in the PICU setting, comparing two modes of invasive mechanical ventilation, PC vs PRVC, using carbon dioxide levels as the outcome measure. We hypothesize that a digital interventional trial comparing two modes of ventilation in the PICU is feasible and that children will spend more time in normocarbia when ventilated using the PRVC mode compared to the PC mode.

## METHODS

### Study design and setting

This study is a pilot, interventional, single-centre, open-label, randomized controlled clinical trial to assess the feasibility of a digital PICU trial. The trial is conducted in the PICU of the University Children’s Hospital Zurich, Switzerland, and is overseen by the Children’s Research Center monitor. This trial serves to evaluate feasibility, test procedures, and obtain parameter estimates that will inform the design and the sample size of future trials.

### Participants, screening and consent procedures

Mechanically ventilated children who are younger than 18 years are eligible for randomization if their weight is above 2 kg, and if they have an arterial line. Patients who have been ventilated for longer than 24 hours, and those with cyanotic heart disease, pulmonary hypertension, or at risk for increased intracranial pressure are excluded. Detailed inclusion and exclusion criteria are listed in Table 1. Mechanically ventilated children are screened daily by study coordinators using the institutional PICU patient data monitoring systems (MetaVision Suite^®^, Israel-iMDsoft Ltd., Tel Aviv, Israel / CGM Clinical – Phoenix^®^, CompuGroup Medical Switzerland AG, Niederwangen, Switzerland). If eligible, a study coordinator and a physician contact the child’s parents or legal guardian(s) to explain the study and obtain prospective consent. Written informed consent is sought from parents or guardians and adolescent patients aged 14 years and older. If prospective consent cannot be obtained for an otherwise eligible child due to the emergency environment, study procedures are initiated if approved by an independent physician and consent is sought as soon as possible once the parent/guardian has had time to adapt to the emergency environment in the PICU (consent to continue, to be sought within 24 hours). No intervention or outcome data will be available for analysis from those where consent cannot be obtained. Patients may be enrolled more than once during subsequent PICU admissions if the date of the second enrolment is at least 30 days after the date of the previous enrolment.

**Table 1.**
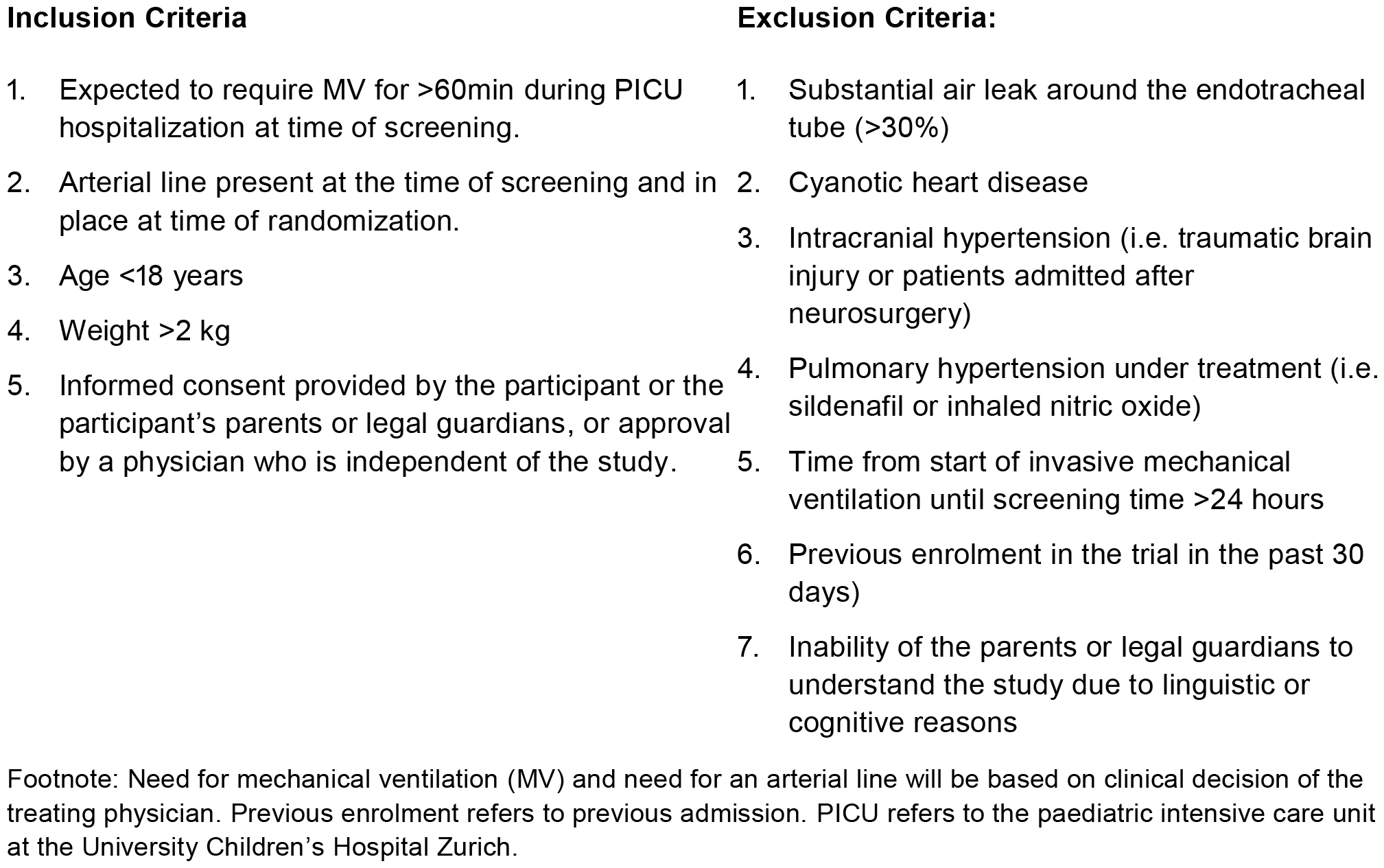
Inclusion and exclusion criteria.

| **Inclusion Criteria** | **Exclusion Criteria:** |
| --- | --- |
| 1. Expected to require MV for >60min during PICU hospitalization at time of screening. | 1. Substantial air leak around the endotracheal tube (>30%) |
| 2. Arterial line present at the time of screening and in place at time of randomization. | 2. Cyanotic heart disease |
| 3. Age <18 years | 3. Intracranial hypertension (i.e. traumatic brain injury or patients admitted after neurosurgery) |
| 4. Weight >2 kg | 4. Pulmonary hypertension under treatment (i.e. sildenafil or inhaled nitric oxide) |
| 5. Informed consent provided by the participant or the participant's parents or legal guardians, or approval by a physician who is independent of the study. | 5. Time from start of invasive mechanical ventilation until screening time >24 hours |
|  | 6. Previous enrolment in the trial in the past 30 days) |
|  | 7. Inability of the parents or legal guardians to understand the study due to linguistic or cognitive reasons |

Footnote: Need for mechanical ventilation (MV) and need for an arterial line will be based on clinical decision of the treating physician. Previous enrolment refers to previous admission. PICU refers to the paediatric intensive care unit at the University Children's Hospital Zurich.

### Randomization and blinding

The randomization schedule was computer-generated with variable block sizes (4, 6, 8 and 10) using a 1:1 allocation ratio and stratified by age as follows:

- neonates, defined as having a postnatal age <28 days if born at term (i.e., born at ≥37 weeks of gestational age) or a postnatal age of <44 weeks of corrected gestational age if born preterm (i.e., born at <37 weeks of gestational age).
- older infants, children, or adolescents, defined as a postnatal age ≥28 days if born at term or ≥44 weeks corrected gestational age if born preterm.

The randomization and electronic allocation system is embedded in the trial’s REDCap database, which ensures concealment of allocation. The randomization sequence was generated by the data manager, who is not involved in recruitment or allocation. Based on our team’s previous experience using REDCap randomisation, we do not anticipate any challenges, and as such, plus the digital focus of the study, we do not have back-up envelopes. Once eligibility is confirmed and consent is obtained from the patient, family or an independent physician, randomization is performed by a study coordinator or physician, and allocation is communicated electronically to the treating staff. Blinding is not possible due to the nature of the intervention. Eligible patients are enrolled in the trial after obtaining consent (time point 0), or consent to continue (Figure 2), and randomization.

**Figure 1.**
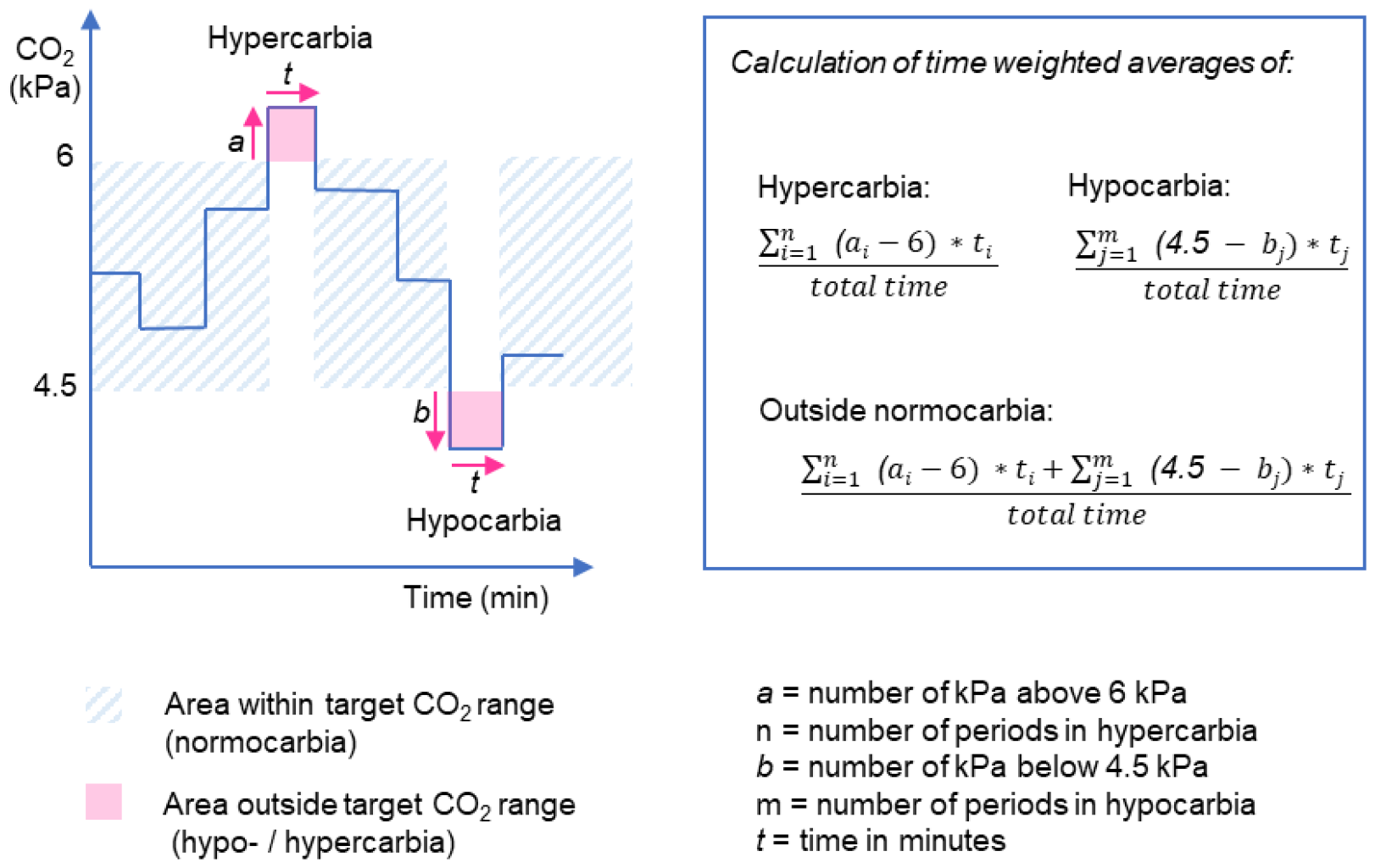
Graphical display and formulas used to calculate time-weighted averages for hypocarbia, hypercarbia and overall time spent outside the CO_2_ target range. The formulas account for both the severity of the hypocarbia / hypercarbia and time spent under hypocarbia / above hypercarbia in the numerator, divided by the total time until censoring for each patient.

**Figure 2.**
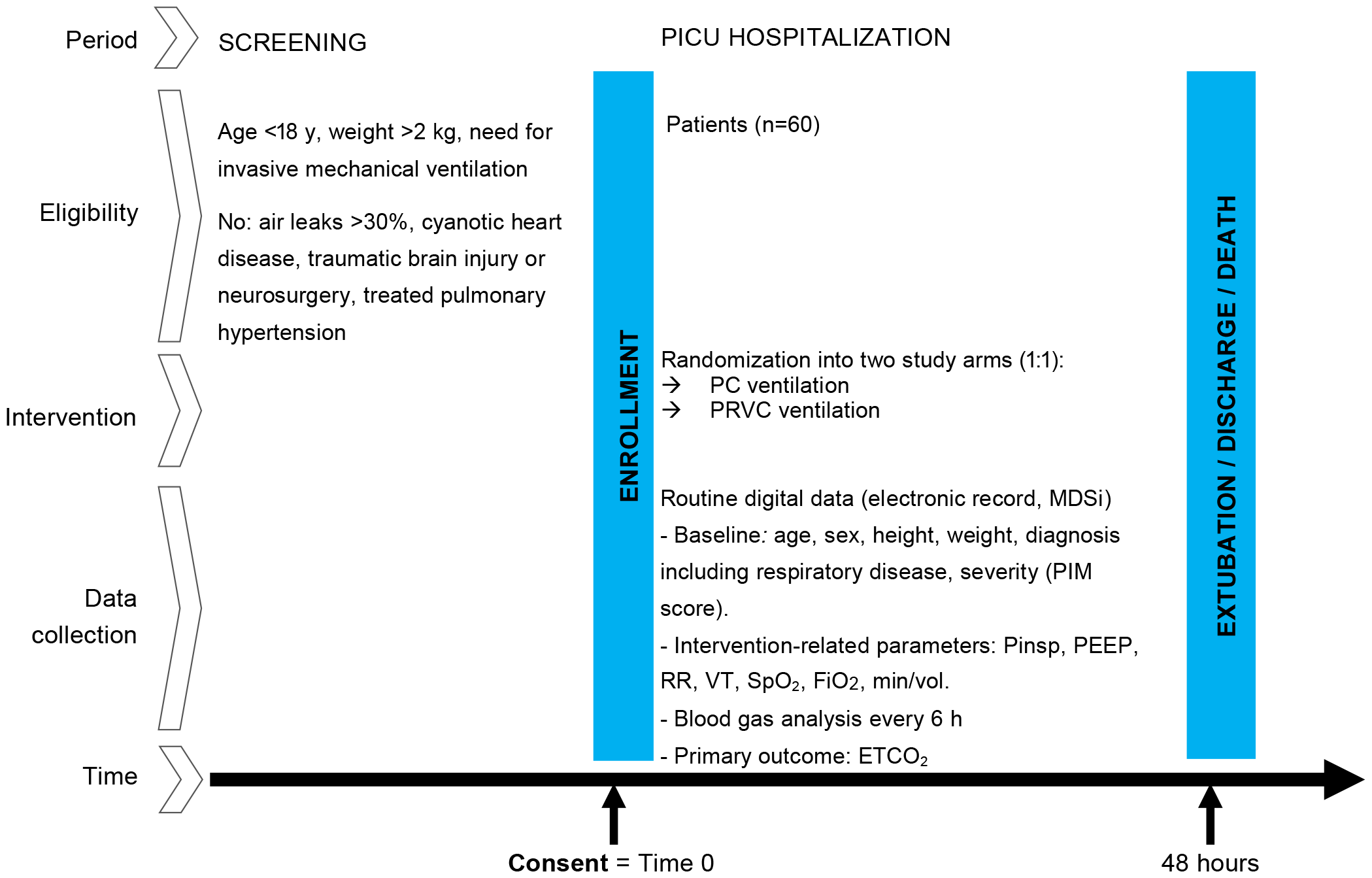
Study design of the CoCO2 randomized controlled trial. Abbreviations: PC = pressure control; PRVC = pressure-regulated volume control; MDSi = minimal dataset of the Swiss Society of Intensive Care, PIM-2 = Paediatric Index of Mortality, Pinsp = peak inspiratory pressure, PEEP = positive end expiratory pressure, RR = respiratory rate, VT =, tidal volume, SpO_2_ = peripheral oxygen saturation, FiO_2_ = Fraction of inhaled oxygen, MV = minute volume, ETCO_2_ = end-tidal partial pressure of carbon dioxide.

### Interventions and treatment arms

The trial compares two ventilation modes: PC and PRVC. Both modes of invasive mechanical ventilation are part of routine standard care at the study hospital. Invasive ventilation is provided using the Bellavista™ 1000 ventilators (IMT Medical, Buchs, Switzerland / Vyaire Medical, Chicago, US). The Bellavista brand label for the PC mode is “pressure support ventilation (PSV)” and for the PRVC mode is “pressure support ventilation with TargetVent (PSV with TargetVent)”. Both ventilation modes provide pressure using a decelerating flow pattern. The difference between the modes is that in PC physicians set the inspiratory pressure (plus inspiratory time, respiratory rate, and positive end-expiratory pressure (PEEP)), whereas in PRVC physicians set a target tidal volume (plus inspiratory time, respiratory rate, PEEP, as well as the upper and lower limits of pressure support), and an algorithm delivers pressure to achieve the target tidal volume based on lung compliance measured during previous breaths while remaining between the prescribed pressure limits. Deviations from the assigned mode are allowed based the clinical judgment of the treating physician and will be captured. Measures to improve adherence include verbal reminders and placement of stickers indicating the assigned ventilation mode on the ventilation monitors.

### Study outcomes

This pilot study is designed to provide estimates of feasibility and physiological outcomes with high temporal resolution to assist in the planning of future trials.

- The *primary feasibility outcome* is adherence to the assigned ventilation mode among enrolled patients, measured from randomization to censoring (48 hours after randomization, extubation, discharge from PICU, or death, whichever comes first) among enrolled patients. This is measured as the proportion of time under invasive ventilation spent in the assigned ventilation mode. Discontinuations from the assigned mode lasting 15 minutes or less are excluded. We set a threshold for adherence to the allocated mode of 80%, meaning that patients who are ventilated with the assigned mode during ≥80% of their time in the study are considered adherent.
- The *primary physiological outcome* is the proportion of time spent within the target CO_2_ range (normocarbia, defined as CO_2_ ≥ 35 mmHg or 4.5 kPa and ≤ 45 mmHg or 6 kPa) as measured by end-tidal CO_2_ (ETCO_2_) recorded by the ventilation device and transferred to the electronic health record system every minute from randomization to censoring. This is calculated as the total number of minutes spent with ETCO_2_ within the target CO_2_ range divided by the total number of minutes ventilated from randomization to censoring. Arterial blood gas analysis is the gold standard to assess ventilation, however this is an invasive and intermittent measurement. Trends in ETCO_2_ are non-invasive, continuously available at bedside, and serve as a proxy for changes in partial pressure of CO_2_ in arterial blood (PaCO_2_) in children and neonates^13^.
- *Secondary feasibility outcomes* describe recruitment and implementation aspects of the study (Table 2). These include the number of patients who were screened, were missed, gave consent, and were randomized and enrolled per month, reasons for protocol violations; and times from randomization to protocol violation, from start of ventilation to screening, from start of ventilation to randomization, and from consent to randomization. The technical feasibility of digital data extraction is assessed through the proportion of enrolled participants with complete primary and secondary outcome data extracted from the electronic patient records.
- *Secondary physiological outcomes* on hypocarbia, hypercarbia, or both are calculated using time-weighted averages to account not only for the time spent outside the target CO_2_ range but also for the magnitude of the difference from the target CO_2_ range (Table 2, Figure 1). The oxygenation index measured using peripheral oxygen saturation (SpO_2_) and arterial oxygen (PaO_2_) are also secondary physiological outcomes. While ETCO_2_, transcutaneous CO_2_, and SpO_2_ are time series measurements, the PaCO_2_ and PaO_2_ are obtained from intermittent blood gas analyses. To assess the later intermittent measurements over study time, each value will be carried on until the time the next measurement^14^.

### Sample size

We aim to recruit a total of 60 patients, 30 per study arm. To determine feasibility to progress to a larger trial, we define feasibility of adherence to the allocated ventilation mode (primary feasibility outcome) if two thirds or more (≥67%) of patients are ventilated with the allocated mode during ≥80% of the time from randomization until censoring. If 47 out of 60 patients are ventilated with the allocated mode for ≥80% of the study time, this would yield a feasibility of adherence of 78% with a one-sided lower 95% confidence interval of 68%. A sample size of 60 participants will allow us to assess other feasibility outcomes, and is considered sufficient to assess the variation and distribution of continuous physiological outcomes for future trials, based on previously published flat^15-18^ and stepped rules of thumb for pilot trials^19^.

**Table 2.**
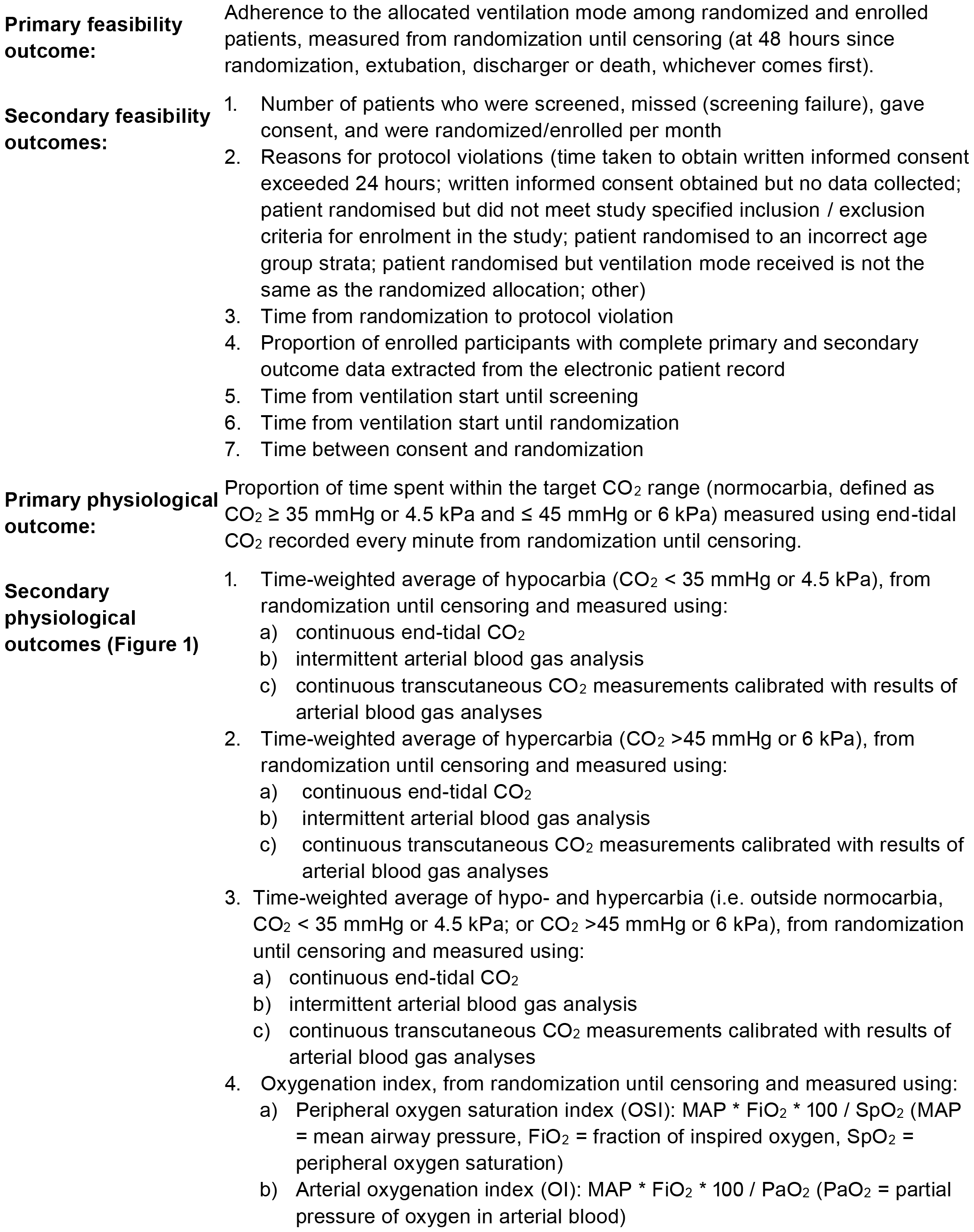
Outcome measures.

### Data collection

There are two primary sources of data for this study: 1) data manually entered into REDCap, and 2) the electronic medical record.

Data for documentation and monitoring purposes that requires manual collection is related to screening, consenting, protocol deviations, and adverse events for reporting and monitoring purposes. This data is recorded using a short electronic Case Report Form (eCRF) in REDCap electronic data capture tools hosted at the University Children’s Hospital Zurich^20^. The data is accessible only to the study team, is password-protected, and backed-up nightly to the servers of the University Children’s Hospital Zurich. This data will be used to calculate a subset of the secondary feasibility outcomes.

Data related to the intervention and physiological outcomes of the trial is captured digitally through the electronic health record. This has two advantages: 1) there is almost no data entry burden for study coordinators, physicians, or nurses on the care team, 2) time series data can be collected at a much higher temporal resolution than what would be possible if done manually. Electronic data sources include: clinical information system (CGM Clinical - Phoenix^®^), intensive care clinical information system (MetaVision^®^), laboratory information system (LIS), and hospital administration system (Hospis^®^). In MetaVision^®^, time series parameters are collected every minute. The following medical devices feed data directly into the intensive care information system: mechanical ventilator (Bellavista^®^) for all ventilation-related parameters and ETCO_2_, transcutaneous CO_2_ monitor (Sentec^®^), SpO_2_ monitor (Pulsoximeter Rad-87 Massimo^®^). The clinical data warehouse team at the University Children’s Hospital Zurich extracts information from the participants’ electronic health record from randomization to censoring as a comma-separated value file. In addition, the mandatory dataset for the Swiss Intensive Care registry (MDSI, https://www.sgi-ssmi.ch/de/datensatz.html) is also captured in the clinical data warehouse and helps describe the patient cohort. The following information is extracted by the clinical data warehouse team to describe the study population, and to calculate the primary feasibility outcome, and the primary and secondary physiological outcomes:

- Baseline characteristics, such as age, sex, height, weight, Paediatric Index of Mortality-2 (PIM-2)^21,22^, which indicates the risk of death as a percentage from 0-100 (the higher the percentage, the higher the risk) and is used as a proxy for overall severity of disease, main diagnosis, date and time of admission, start of ventilation and randomization, arterial blood gas analysis, endotracheal tube size, and cuff.
- Respiratory mechanics and intervention-related parameters during the study period, including PEEP, mean airway pressure (MAP), peak inspiratory pressure (PIP), respiratory rate, tidal volume, minute volume, inspiratory time, leakage percentage, fraction of inspired oxygen (FiO_2_), and SpO_2_
- Outcome-related parameters during the study period such as ventilation mode, ETCO_2_, transcutaneous CO_2_, date and time of extubation, discharge or death, inspired fraction of oxygen ratio (SpO_2_/FiO_2_), results of consecutive blood gas analyses and arterial partial pressure of oxygen and inspired fraction of oxygen ratio (PaO_2_/FiO_2_). Arterial blood gas analyses are planned to be obtained every 6 hours during the study period. If the arterial line is removed during the study period, blood gas analyses may be obtained preferably from a capillary sample.

To test the electronic patient data extraction procedures, we performed test extractions of the above parameters using retrospective data from the electronic health records of 60 patients with approved general consent. This allowed us to streamline data extraction and cleaning procedures before recruitment started. In addition, we will perform a manual data verification to validate the data extraction after enrolment of the first 2 to 10 patients.

### Statistical analysis plan

The primary analysis will be performed on an intention-to-treat cohort. In case patients participate in the trial multiple times, the unit of analysis will shift from individual patients to individual admissions. This distinction will be appropriately handled through multilevel analyses. Results will be presented as effect estimates and 95% confidence intervals, without the inclusion of p-values. While missing data imputation will not be performed, outliers management in time series data will be addressed through the application of kernel smoothing^23^.

We will use descriptive statistics to summarize patient characteristics, adverse events, and the distribution of feasibility and physiological outcomes. Categorical variables will be described using frequencies and percentages, continuous variables following a normal distribution using mean and standard deviation, and non-normally distributed variables using median and interquartile range. Normality will be assessed graphically with plots (histograms and Q-Q plots).

The analysis of both primary and secondary feasibility outcomes will be descriptive.

To analyse the primary physiological outcome, we will calculate the mean difference in the proportion of time spent in normocarbia (outcome) between the two ventilation modes (explanatory variable) using a linear mixed-effects regression model if the outcome is normally distributed. The stratification variable (age group) will be included as a fixed effect, and admissions will be nested within patients to account for correlated observations from patients potentially included more than once in the study. If the outcome has a skewed distribution, we will use quantile regression, also adjusting for stratification. The study is not powered to detect differences in physiological outcomes between the two ventilation models. Secondary physiological outcomes will be calculated using time-weighted averages as described in Figure 1.

For physiological outcomes, we will conduct a per-protocol analysis including patients with adherence to the assigned ventilation mode of 80% or more of the study time.

As sensitivity analysis, we will calculate physiological outcomes using individual target CO_2_ ranges manually entered into the electronic information system by prescribing physicians when available, instead of standard normocarbia ranges. We will also assess the robustness of the results by testing different kernel sizes for smoothing and also present crude (not smoothed) results.

Secondary analyses will explore triggers and factors associated with deviations from the assigned ventilation mode (if patients received a mechanical ventilation mode different from that assigned at randomization). We will examine interactions with age for physiological outcomes using plots and likelihood ratio tests. We will assess agreement between PaCO_2_ and both ETCO_2_ and transcutaneous CO_2_ values around the time windows when blood gas analysis was performed.

### Ethics

This study has received ethical approval (ethics committee of the canton of Zurich, BASEC 2022-00829). Substantial changes to the study setup and study organization, the protocol and relevant study documents will be submitted to the Ethics Committee for approval before implementation. Trial and participant data are only accessible to authorized personnel who need the data to fulfil their duties within the scope of the study. Participants are coded with a unique participant identifier on the CRFs and other study specific documents. Coding documents linking the participant personal information to unique study identifiers are kept in a password-protected Excel spreadsheet with limited access.

### Dissemination

Study results will be disseminated using traditional (submission for publication in a peer-reviewed journal and presentations at conferences) and novel (social media, web, or podcasts) formats. Information about the study is available on the PICU page of the University Children’s Hospital Zurich – Children’s Research Centre webpage (https://www.kispi.uzh.ch/forschungszentrum/forschungsgebiete/intensivmedizin-neonatologie-fzk). Participants are asked during the consent process if they would like to be informed of the results of this study and, if so, their email address is collected for the purpose of sharing relevant publications.

### Patient and public involvement

Patient and public involvement has not been implemented in this pilot study, which focuses on feasibility and mechanistic aspects rather than translation to clinical care at this stage. We fully recognize the importance of involving patient families in the early stages of the research plan. If this pilot trial is successful, we plan to involve patient representatives in the design of future larger follow-up trials, with a stronger focus on patient-centred outcomes and clinical implications.

### Monitoring and safety

Monitoring is performed according to the pre-defined monitoring plan of the Children’s Research Center of the University Children’s Hospital Zurich. The site initiation visit took place prior to the enrolment of any participants. We plan three routine onsite monitoring visits (1^st^ as soon as possible after the enrolment of the first two participants, 2^nd^ after enrolment of 30 participants, 3^rd^ and close-out visit after enrolment of the last participant or in case of premature closure). The monitor reviews all informed consent forms, and at least one complete review of the trial master file / investigator site file. Eligibility, primary endpoint, serious adverse events and protocol-specific safety parameters are reviewed for the first two trial participants and up to 20% of trial participants enrolled at the time of the visit, if available. Randomization and allocation will be reviewed for at least 20% of trial participants.

The study design does not pose any serious additional risks to patients, as patients require the use of invasive mechanical ventilation due to their clinical situation, and both modes of invasive mechanical ventilation are part of accepted standard of care at the study site. Exemptions from expedited reporting are allowed when the serious adverse event is either a clear result of the underlying condition or a known complication of the condition. Because both interventions are currently used in paediatric critical clinical care, the following serious adverse events that may occur in mechanically ventilated critically ill patients are exempt from expedited reporting: pneumothorax, endotracheal tube obstruction, pneumonia, sepsis, cerebral oedema, seizures, stroke, intraventricular haemorrhage, cardiac failure, atelectasis, air leaks, pulmonary haemorrhage, subglottic stenosis, vocal cord injury, and need for extracorporeal membrane oxygenation (ECMO). Deaths cases will be reported. We will submit an annual safety report to the ethics committee. Because this is a pilot feasibility trial investigating two currently accepted and commonly used ventilation modes with high anticipated safety profile and minimal associated risk, with main focus on digital trial data extraction procedures, a formal data safety monitoring board was not considered necessary.

## TRIAL STATUS AND TIMELINE

The recruitment period started on 15^th^ of January 2024 and the estimated duration of the recruitment period is 6 months.

## SIGNIFICANCE

The richness and high resolution of data routinely collected in intensive care provides a unique opportunity to study the efficacy of different procedures and treatments on physiological outcomes in children, and can enable more efficient digital trial designs leveraging data extraction procedures to reduce workload associated with trials. This study serves as proof of concept for future larger digital clinical trials with patient-centred outcomes using electronic health record data. This data can assist in the design of future trials by providing necessary information for sample size calculation and by identifying potential difficulties and solutions.

## Supporting information

Statistical analysis plan

SPIRIT checklist

## AUTHOR CONTRIBUTIONS

RM, LJS and JB conceptualized the study and obtained funding. RM drafted the first version of this manuscript. All authors contributed to the study design and/or contribute to study implementation. KG provided statistical expertise and DC will conduct statistical analysis. BBa and VJ designed and implement electronic health record data extraction pipelines for data collection. DC and RM supported assessment and validation of the data extraction pipeline. All authors critically reviewed and approved the manuscript.

## DATA AVAILABILITY

The research team will have access to the dataset of this study. Anonymized data could be available upon reasonable request after approval from the research committee and sponsoring institution.

## AKNOWLEGMENTS

We thank patients and families for their participation in this study, the PICU team of the University Children’s Hospital Zurich team for their support during study conduct, Sanella Ellersich and Elisa Zimmerman for their support in study coordination and management, Melanie Huber for her support in database and data management.

## APPENDICES

Statistical analysis plan

Informed consent form

SPIRIT checklist

## ADMINISTRATIVE INFORMATION

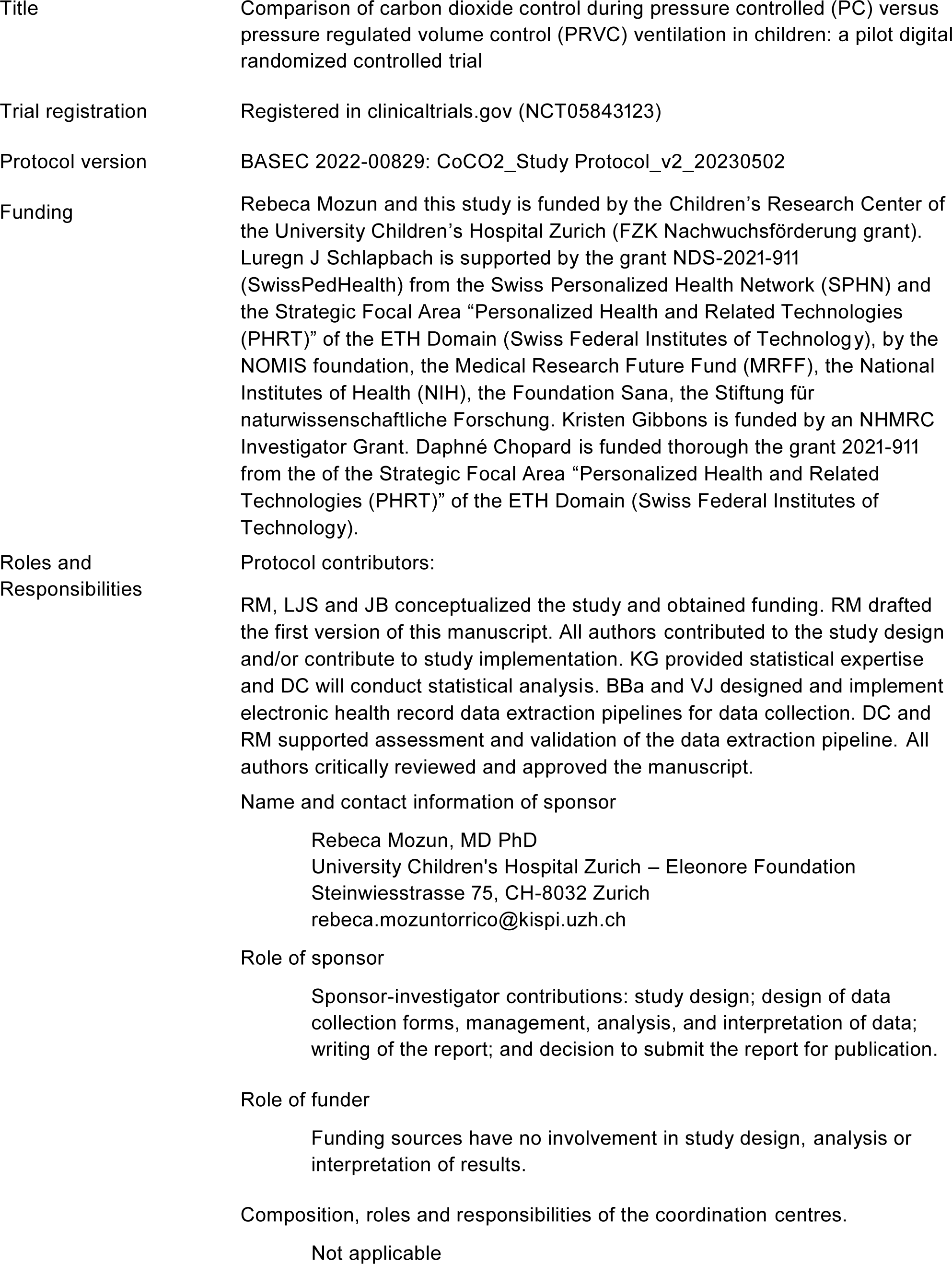

|  |  |
| --- | --- |
| Title | Comparison of carbon dioxide control during pressure controlled (PC) versus pressure regulated volume control (PRVC) ventilation in children: a pilot digital randomized controlled trial |
| Trial registration | Registered in clinicaltrials.gov (NCT05843123) |
| Protocol version | BASEC 2022-00829: CoCO2_Study Protocol_v2_20230502 |
| Funding | Rebeca Mozun and this study is funded by the Children's Research Center of the University Children's Hospital Zurich (FZK Nachwuchsförderung grant). Luregn J Schlapbach is supported by the grant NDS-2021-911 (SwissPedHealth) from the Swiss Personalized Health Network (SPHN) and the Strategic Focal Area "Personalized Health and Related Technologies (PHRT)" of the ETH Domain (Swiss Federal Institutes of Technology), by the NOMIS foundation, the Medical Research Future Fund (MRFF), the National Institutes of Health (NIH), the Foundation Sana, the Stiftung für naturwissenschaftliche Forschung. Kristen Gibbons is funded by an NHMRC Investigator Grant. Daphné Chopard is funded thorough the grant 2021-911 from the of the Strategic Focal Area "Personalized Health and Related Technologies (PHRT)" of the ETH Domain (Swiss Federal Institutes of Technology). |
| Roles and Responsibilities | Protocol contributors:<br>RM, LJS and JB conceptualized the study and obtained funding. RM drafted the first version of this manuscript. All authors contributed to the study design and/or contribute to study implementation. KG provided statistical expertise and DC will conduct statistical analysis. BBa and VJ designed and implement electronic health record data extraction pipelines for data collection. DC and RM supported assessment and validation of the data extraction pipeline. All authors critically reviewed and approved the manuscript.<br>Name and contact information of sponsor<br>Rebeca Mozun, MD PhD<br>University Children's Hospital Zurich – Eleonore Foundation<br>Steinwiesstrasse 75, CH-8032 Zurich<br>Role of sponsor<br>Sponsor-investigator contributions: study design; design of data collection forms, management, analysis, and interpretation of data; writing of the report; and decision to submit the report for publication.<br>Role of funder<br>Funding sources have no involvement in study design, analysis or interpretation of results.<br>Composition, roles and responsibilities of the coordination centres.<br>Not applicable |

## REFERENCES

1. Arabi YM, Azoulay E, Al-Dorzi HM, et al. How the COVID-19 pandemic will change the future of critical care. Intensive Care Medicine 2021; 47(3): 282–91.

2. Wise J, Coombes R. Covid-19: The inside story of the RECOVERY trial. BMJ 2020; 370: m2670.

3. Inan OT, Tenaerts P, Prindiville SA, et al. Digitizing clinical trials. NPJ Digit Med 2020; 3: 101.

4. Semler MW, Casey JD, Lloyd BD, et al. Oxygen-Saturation Targets for Critically Ill Adults Receiving Mechanical Ventilation. N Engl J Med 2022; 387(19): 1759–69.

5. Khemani RG, Markovitz BP, Curley MAQ. Characteristics of Children Intubated and Mechanically Ventilated in 16 PICUs. Chest 2009; 136(3): 765–71.

6. Jauncey-Cooke JI, Bogossian F, East CE. Lung protective ventilation strategies in paediatrics-A review. Aust Crit Care 2010; 23(2): 81–8.

7. Belteki G, Morley CJ. Volume-Targeted Ventilation. Clin Perinatol 2021; 48(4): 825–41.

8. Abu-Sultaneh S, Iyer NP, Fernández A, et al. Executive Summary: International Clinical Practice Guidelines for Pediatric Ventilator Liberation, A Pediatric Acute Lung Injury and Sepsis Investigators (PALISI) Network Document. American Journal of Respiratory and Critical Care Medicine 2023; 207(1): 17–28.

9. Kneyber MCJ, de Luca D, Calderini E, et al. Recommendations for mechanical ventilation of critically ill children from the Paediatric Mechanical Ventilation Consensus Conference (PEMVECC). Intensive Care Med 2017; 43(12): 1764–80.

10. Platen PV, Pomprapa A, Lachmann B, Leonhardt S. The dawn of physiological closed-loop ventilation-a review. Crit Care 2020; 24(1): 121.

11. Klingenberg C, Wheeler KI, McCallion N, Morley CJ, Davis PG. Volume-targeted versus pressure-limited ventilation in neonates. Cochrane Database Syst Rev 2017; 10: CD003666.

12. Emeriaud G, López-Fernández YM, Iyer NP, et al. Executive Summary of the Second International Guidelines for the Diagnosis and Management of Pediatric Acute Respiratory Distress Syndrome (PALICC-2). Pediatr Crit Care Med 2023; 24(2): 143–68.

13. McDonald MJ, Montgomery VL, Cerrito PB, Parrish CJ, Boland KA, Sullivan JE. Comparison of end-tidal CO2 and Paco2 in children receiving mechanical ventilation. Pediatric Critical Care Medicine 2002; 3(3): 244–9.

14. Fabres J, Carlo WA, Phillips V, Howard G, Ambalavanan N. Both Extremes of Arterial Carbon Dioxide Pressure and the Magnitude of Fluctuations in Arterial Carbon Dioxide Pressure Are Associated With Severe Intraventricular Hemorrhage in Preterm Infants. Pediatrics 2007; 119(2): 299–305.

15. Browne RH. On the use of a pilot sample for sample size determination. Stat Med 1995; 14(17): 1933–40.

16. Julious SA. Sample size of 12 per group rule of thumb for a pilot study. Pharmaceutical Statistics 2005; 4(4): 287–91.

17. Kieser M, Wassmer G. On the Use of the Upper Confidence Limit for the Variance from a Pilot Sample for Sample Size Determination. Biometrical Journal 1996; 38: 941–9.

18. Sim J, Lewis M. The size of a pilot study for a clinical trial should be calculated in relation to considerations of precision and efficiency. J Clin Epidemiol 2012; 65(3): 301–8.

19. Whitehead AL, Julious SA, Cooper CL, Campbell MJ. Estimating the sample size for a pilot randomised trial to minimise the overall trial sample size for the external pilot and main trial for a continuous outcome variable. Stat Methods Med Res 2016; 25(3): 1057–73.

20. Harris PA, Taylor R, Thielke R, Payne J, Gonzalez N, Conde JG. Research electronic data capture (REDCap)—A metadata-driven methodology and workflow process for providing translational research informatics support. Journal of Biomedical Informatics 2009; 42(2): 377–81.

21. Shann F, Pearson G, Slater A, Wilkinson K. Paediatric index of mortality (PIM): a mortality prediction model for children in intensive care. Intensive Care Med 1997; 23(2): 201–7.

22. Slater A, Shann F, Pearson G, Paediatric Index of Mortality Study G. PIM2: a revised version of the Paediatric Index of Mortality. Intensive Care Med 2003; 29(2): 278–85.

23. Härdle W, Vieu P. KERNEL REGRESSION SMOOTHING OF TIME SERIES. Journal of Time Series Analysis 1992; 13(3): 209–32.

