## Supplementary material for "Comparison of carbon dioxide control during pressure controlled versus pressure regulated volume controlled ventilation in children (CoCO2): protocol for a pilot digital randomized controlled trial": Statistical analysis plan

**Acronym:** CoCO2

**Trial registration number:** NCT05843123

**Statistical analysis plan version:** 1.0, 27.03.2024

**Protocol version:** v2, 02.05.2023

**Statistical analysis plan revision history:**

| Version, date | Revision | Reason for revision | Timing of revision in relation to interim analysis |
|---|---|---|---|

**TABLE OF CONTENTS:**

**Roles and responsibilities** 2
**Introduction** 3
**Study Methods** 3
Statistical principals 4
Trial Population 4
Analysis 5
**References** **Error! Bookmark not defined.**

### Roles and responsibilities

| Name | Affiliation | Data related roles and responsibilities |
|---|---|---|
| Rebeca Mozun | Department of Intensive Care and Neonatology and Children's Research Centre, University Children's Hospital Zurich, University of Zurich, Zurich, Switzerland | *Role*: Principal investigator<br>*Responsibly*:<br>• set-up REDCap database<br>• implementation of risk based quality management system<br>• support data validation and statistical analyses<br>• reporting |
| Daphné Chopard | Department of Intensive Care and Neonatology and Children's Research Centre, University Children's Hospital Zurich, University of Zurich, Zurich, Switzerland<br><br>Department of Computer Science, ETH Zurich, Zurich, Switzerland | *Role:* Postdoctoral researcher<br>*Responsibility*<br>• support data validation<br>• perform statistical analyses |
| Kristen Gibbons | Child Health Research Centre, The University of Queensland, Brisbane, Australia | *Role*: Statistics and epidemiology expert<br>*Responsibility*<br>• provide input on protocol, analysis plan<br>• supervise methodology and statistical analysis |
| Beat Bangerter | IT Department and Children's Research Centre, University Children's Hospital Zurich, University of Zurich, Zurich, Switzerland | Role: Lead data intelligence group<br>*Responsibility*:<br>• supervise development of data extraction pipeline |
| Vera Jäggi | IT Department and Children's Research Centre, University Children's Hospital Zurich, University of Zurich, Zurich, Switzerland | *Role*: Senior data engineer<br>*Responsibility*:<br>• develop data extraction pipeline<br>• extract data from clinical electronic records |
| Melanie Huber | Department of Intensive Care and Neonatology and Children's Research Centre, University Children's Hospital Zurich, University of Zurich, Zurich, Switzerland | *Role*: Data manager<br>*Responsibility*:<br>• support redcap database creation and exports, maintenance, data validation |
| Anika Adam & Sanella Ellersich | Department of Intensive Care and Neonatology and Children's Research Centre, University Children's Hospital Zurich, University of Zurich, Zurich, Switzerland | *Role*: PINNACLE study coordinator<br>*Responsibility*:<br>• screen and recruit participants<br>• enter data in REDCap<br>• documentation (study folder, log files)<br>• data validation |
| Angela Beccarelli | Children's Research Centre, University Children's Hospital Zurich, University of Zurich, Zurich, Switzerland | *Role*: monitor<br>*Responsibility*:<br>• trial monitoring audits |

_
Dr. Rebeca Mozun
Sponsor-investigator
University Children's Hospital Zurich
Date:

Prof. Kristen Gibbons
Senior statistician
University of Queensland
Date:

### Introduction

#### Background and rationale

Trials using digital recordings of routine clinical data have great potential to decrease the resources required to conduct clinical trials, which remain one of the major obstacles to faster evidence generation in clinical care. Intensive care units (ICUs) represent an attractive setting for digital trials given the rich data environment with high temporal resolution, yet these are scarcely done.

The randomization and electronic allocation system is embedded in the trial's REDCap database, which ensures concealment of allocation. Blinding of the intervention is not possible due to the nature of the intervention.

#### Sample size

We aim to recruit a total of 60 patients, 30 per study arm. To determine feasibility to progress to a larger trial, we define feasibility of adherence to the allocated ventilation mode (primary feasibility outcome) if two thirds or more (≥67%) of patients are ventilated with the allocated mode during ≥80% of the time from randomization until censoring. If 47 out of 60 patients are ventilated with the allocated mode for ≥80% of the study time, this would yield a feasibility of adherence of 78% with a one-sided lower 95% confidence interval of 68%. A sample size of 60 participants will allow us to assess other feasibility outcomes, and is considered sufficient to assess the variation and distribution of continuous physiological outcomes for future trials, based on previously published flat and stepped rules of thumb for pilot trials.

#### Framework

The main aim of this trial is to test feasibility, not to prove superiority. We will test procedures and to obtain parameter estimates that will help plan and sample size calculations of future larger trials.

#### Statistical interim analysis and stopping guidance

No interim analysis is planned and there are no stopping rules.

We will perform a manual data verification to validate the data extraction after enrolment of the first 2 to 10 patients.

#### Timing of final analysis

Final analysis will be performed after all data endpoints are collected and monitoring activities are completed for all participants (n=60).

#### Timing of outcome assessments

Given the digital components, data on the endpoints will be collected in real time using monitoring equipment, with a goal of one extraction per week.

### Statistical principals

#### Confidence intervals and p values

Results will be presented as effect estimates and 95% confidence intervals, without the inclusion of p-values, for all primary and secondary outcomes.

#### Adherence and protocol deviations

Our primary feasibility outcome is adherence to the assigned ventilation mode among enrolled patients, measured from randomization to censoring (48 hours after randomization, extubation, discharge from PICU, or death, whichever comes first) among enrolled patients. This is measured as the proportion of time under invasive ventilation spent in the assigned ventilation mode. Discontinuations from the assigned mode lasting 15 minutes or less are excluded. We set a threshold for adherence to the allocated mode of 80%, meaning that patients who are ventilated with the assigned mode during ≥80% of their time in the study are considered adherent.

We will record the following reasons for protocol violations: time taken to obtain written informed consent exceeded 24 hours; written informed consent obtained but no data collected; patient randomised but did not meet study specified inclusion / exclusion criteria for enrolment in the study; patient randomised to an incorrect age group strata; patient randomised but ventilation mode received is not the same as the randomized allocation; other).

#### Analysis populations

Main analysis will be done on an intention to treat basis.

For physiological outcomes, we will conduct a per-protocol analysis including patients with adherence to the assigned ventilation mode of 80% or more of the study time.

### Trial Population

#### Screening data

Mechanically ventilated children are screened daily on weekdays by study coordinators using the institutional PICU patient data monitoring systems.

#### Eligibility

Inclusion Criteria:

- Admission to PICU at the University Children's Hospital Zurich
- Need for mechanical ventilation for >60 min during PICU hospitalization. Need for mechanical ventilation will be based on clinical decision of the treating physician.
- Need for an arterial line during PICU hospitalization, including at the time of screening and randomization. Need for an arterial line will be based on clinical decision of the treating physician.
- Age <18 years
- Weight >2 kg
- Informed consent provided by the participant or the participant's parents or legal guardians. In case of an emergency situation, a physician who is independent of the research project must be consulted prior to inclusion in order to safeguard the interests of the test subject.

Exclusion Criteria:

- Substantial air leaks around the endotracheal tube (>30%)
- Cyanotic heart disease
- Intracranial hypertension (i.e. traumatic brain injury or patients admitted after neurosurgery)
- Pulmonary hypertension under treatment (i.e. sildenafil or inhaled nitric oxide)
- Time span between ventilation start and randomization >24 hours

- Previous enrolment in the trial in the past 30 days
- Inability of the parents or legal guardians to understand the study due to linguistic or cognitive reasons

#### Recruitment

If eligible, a study coordinator and a physician contact the child's parents or legal guardian(s) to explain the study and obtain prospective consent. Written informed consent is sought from parents or guardians and adolescent patients aged 14 years and older. If prospective consent cannot be obtained for an otherwise eligible child due to the emergency environment, study procedures are initiated if approved by an independent physician and consent is sought as soon as possible once the parent/guardian has had time to adapt to the emergency environment in the PICU (consent to continue, to be sought within 24 hours).

#### Withdrawal / Follow-up

In case of withdrawal of informed consent, there will be no further data collection. All data collected up to that point will be analysed in coded form and anonymized after the evaluation.

There is no planned follow-up of participants.

#### Baseline patient characteristics

We will describe age, sex, height, weight, Paediatric Index of Mortality-2 (PIM-2), which indicates the risk of death as a percentage from 0-100 (the higher the percentage, the higher the risk) and is used as a proxy for overall severity of disease, main diagnosis, date and time of admission, start of ventilation and randomization, arterial blood gas analysis, endotracheal tube size, and cuff.

### Analysis

#### Outcome definitions

Primary feasibility outcome:

The primary feasibility outcome is adherence to the assigned ventilation mode among enrolled patients, measured from randomization to censoring (48 hours after randomization, extubation, discharge from PICU, or death, whichever comes first) among enrolled patients. This is measured as the proportion of time under invasive ventilation spent in the assigned ventilation mode. Discontinuations from the assigned mode lasting 15 minutes or less are excluded. We set a threshold for adherence to the allocated mode of 80%, meaning that patients who are ventilated with the assigned mode during ≥80% of their time in the study are considered adherent.

Secondary feasibility outcomes:

1. Number of patients who were screened, missed (screening failure), gave consent, and were randomized/enrolled per month
2. Reasons for protocol violations (time taken to obtain written informed consent exceeded 24 hours; written informed consent obtained but no data collected; patient randomised but did not meet study specified inclusion / exclusion criteria for enrolment in the study; patient randomised to an incorrect age group strata; patient randomised but ventilation mode received is not the same as the randomized allocation; other)
3. Time from randomization to protocol violation
4. Proportion of enrolled participants with complete primary and secondary outcome data extracted from the electronic patient record
5. Time from ventilation start until screening
6. Time from ventilation start until randomization
7. Time between consent and randomization

Primary physiological outcome:

The primary physiological outcome is the proportion of time spent within the target $CO_2$ range (normocarbia, defined as $CO_2 \geq 35$ mmHg or 4.5 kPa and $\leq 45$ mmHg or 6 kPa) as measured by end-tidal $CO_2$ ($ETCO_2$) recorded by the ventilation device and transferred to the electronic health record system every minute from randomization to censoring. This is calculated as the total number of minutes spent with $ETCO_2$ within the target $CO_2$ range divided by the total number of minutes ventilated from randomization to censoring.

Secondary physiological outcomes (Figure 1)

1. Time-weighted average of hypocarbia ($CO_2 < 35$ mmHg or 4.5 kPa), from randomization until censoring and measured using:
    a. continuous end-tidal $CO_2$
    b. intermittent arterial blood gas analysis
    c. continuous transcutaneous $CO_2$ measurements calibrated with results of arterial blood gas analyses

2. Time-weighted average of hypercarbia (CO2 >45 mmHg or 6 kPa), from randomization until censoring and measured using:
    a. continuous end-tidal CO2
    b. intermittent arterial blood gas analysis
    c. continuous transcutaneous CO2 measurements calibrated with results of arterial blood gas analyses
3. Time-weighted average of hypo- and hypercarbia (i.e. outside normocarbia, CO2 < 35 mmHg or 4.5 kPa; or CO2 >45 mmHg or 6 kPa), from randomization until censoring and measured using:
    a. continuous end-tidal CO2
    b. intermittent arterial blood gas analysis
    c. continuous transcutaneous CO2 measurements calibrated with results of arterial blood gas analyses
4. Oxygenation index, from randomization until censoring and measured using:
    a. Peripheral oxygen saturation index (OSI): MAP * FiO2 * 100 / SpO2 (MAP = mean airway pressure, FiO2 = fraction of inspired oxygen, SpO2 = peripheral oxygen saturation)
    b. Arterial oxygenation index (OI): MAP * FiO2 * 100 / PaO2 (PaO2 = partial pressure of oxygen in arterial blood)

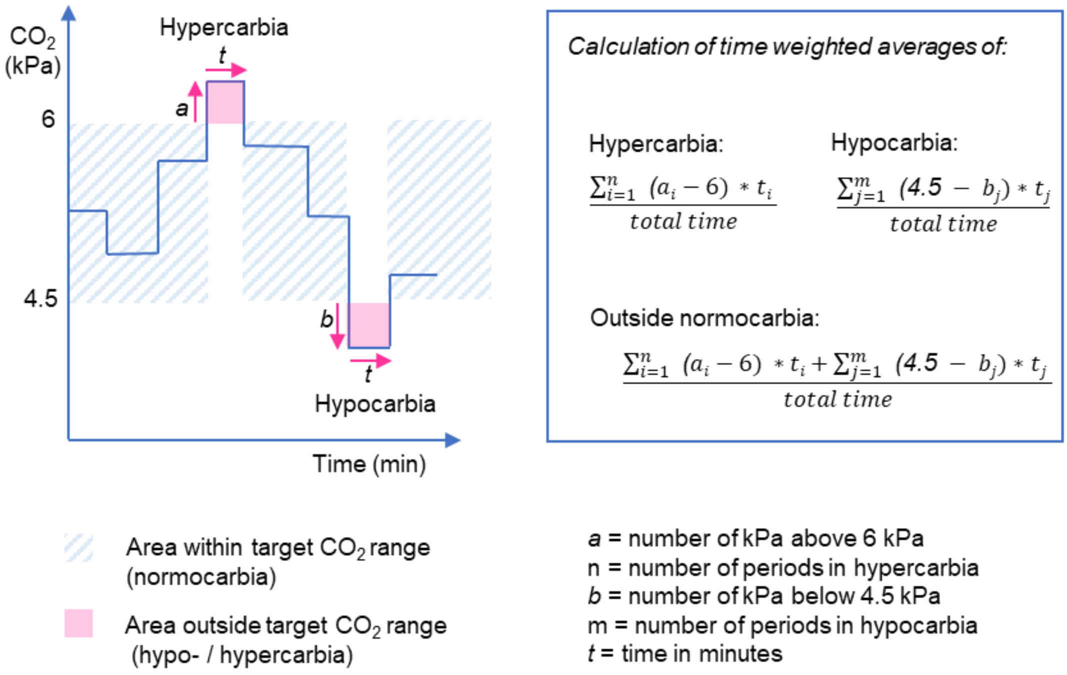

**Figure 1:** Graphical display and formulas used to calculate time-weighted averages for hypocarbia, hypercarbia and overall outside the target range of CO2.
The formulas show that we account for both the severity of the hypocarbia / hypercarbia and time spent under hypocarbia / above hypercarbia in the numerator, and we divide by the total time until censoring for each patient.

### Analysis methods

The primary analysis will be performed on an intention-to-treat cohort. In case patients participate in the trial multiple times, the unit of analysis will shift from individual patients to individual admissions. This distinction will be appropriately handled through multilevel analyses.

For physiological outcomes, we will conduct a per-protocol analysis including patients with adherence to the assigned ventilation mode of 80% or more of the study time.

We will use descriptive statistics to summarize patient characteristics, adverse events, and the distribution of feasibility and physiological outcomes. Categorical variables will be described using frequencies and percentages, continuous variables following a normal distribution using mean and standard deviation, and non-normally distributed variables using median and interquartile range. Normality will be assessed graphically with plots (histograms and Q-Q plots).

### Missing data

While missing data imputation will not be performed, outliers management in time series data will be addressed through the application of kernel smoothing. As data extraction will be digital (direct extraction from electronic patient records), we expect a low proportion of missing values, and if present, they are likely to be due to technical failure.

### Additional analysis

As sensitivity analysis, we will calculate physiological outcomes using individual target CO2 ranges manually entered into the electronic information system by prescribing physicians when available, instead of standard normocarbia ranges. We will also assess the robustness of the results by testing different kernel sizes for smoothing and also present crude (not smoothed) results.

Secondary analyses will explore triggers and factors associated with deviations from the assigned ventilation mode (if patients received a mechanical ventilation mode different from that assigned at randomization). We will examine interactions with age for physiological outcomes using plots and likelihood ratio tests. We will assess agreement between PaCO2 and both ETCO2 and transcutaneous CO2 values around the time windows when blood gas analysis was performed.

To test the electronic patient data extraction procedures, we performed test extractions of the above parameters using retrospective data from the electronic health records of 60 patients with approved general consent. This allowed us to streamline data extraction and cleaning procedures before recruitment started.

### Harms

The study design does not pose any serious additional risks to patients, as patients require the use of invasive mechanical ventilation due to their clinical situation, and both modes of invasive mechanical ventilation are part of accepted standard of care at the study site.

We will manage data with utmost attention to discretion and security, but we acknowledge that the use of digital health information for research carries a risk of data leakage or mishandling. Trial and participant data will only be accessible to authorized personnel who require the data to fulfil their duties within the scope of the study. On the data collection form and other study-specific documents, participants are only identified by a unique participant number. Any document linking the subject number to subject identifiers will be kept in a password-protected Excel sheet with limited access.

### Statistical software

Python, version 3.7, Python Software Foundation. Stata, version 17.0, StataCorp, Texas, USA.
